# COVID-19: Estimation of the Actual Onset of Local Epidemic Cycles, Determination of Total Number of Infective, and Duration of the Incubation Period

**DOI:** 10.1101/2020.08.23.20180356

**Authors:** Rogerio Atem de Carvalho, Eduardo Atem de Carvalho

**Affiliations:** B.Sc. In Informatics, M.Sc., D.Sc. In Production Engineering, Innovation Hub, Instituto Federal Fluminense, Brazil; B.Sc., M.Sc., PhD in Mechanical Engineering, Advanced Materials Laboratory, Universidade Estadual do Norte Fluminense, Brazil

**Keywords:** COVID-19, Pandemics, Infection Control, Longitudinal Studies, Statistical Modeling, Epidemic Cycles

## Abstract

**Background:** Most studies of the epidemic cycles of the pandemic of Sars-CoV-2, or COVID-19 as it became known, define the beginning of specific cycles in countries from the laboratory identification of the first cases of infection, however, there is the awareness that cycles may have started earlier, without proper identification. This influences all the parameters that govern the statistical models used for controlling the infection.

**Purpose:** This work proposes two models based on experimental data. The Logistic Model it is used to obtain three parameters of the epidemic cycle of COVID-19, namely: the final count for the total infected, the daily infection rate and the lag time. Complimentary, a novel inventory model is proposed to calculate the number of infective persons, as well as to determine the incubation period.

**Methods:** The data on epidemic cycles of Germany, Italy, and Sweden are treated previously by the Moving Average Method with Initial value (MAMI), then a variation of the Logistic Model, obtained through curve-fitting, is used to obtain the three parameters. The inventory model is introduced to calculate the actual number of infected persons and the behavior of the incubation period is analyzed.

**Results:** After comparing data from the three countries it is possible to determine the actual probable dates of the beginning of the epidemic cycles for each one, determine the size of the incubation period, as well as to determine the total number of infective persons during the cycle.

**Conclusions:** The actual probable dates of the beginning of the epidemic cycles in the countries analyzed are determined, the total number of infected is determined, and it is statistically proven that the incubation cycle for Sars-CoV-2 is five days.

## 1. Introduction

The analysis of the life cycles of any epidemic involves the analysis of a series of quantitative parameters that govern these cycles and which, given the inherent uncertainty of these events, are generally treated by statistical models. For a number of practical reasons registration of deaths and of infections are inevitably imprecise, although can be corrected over time, as we discuss in [1]. This previous work suggests that one way around this effect is to apply the Moving Average Method with Initial Value (MAMI). In the series analysis, the MAMI is seen as a moving window in time, starting from the first event (infection or death) to the last one. While in [1] we dealt with finding patterns in the number of deaths cycles, in order to predict the end of these cycles, in [2] we introduce a method to estimate the transmission rate or, in technical terms, the Basic Reproduction Number, which in fact, we consider that varies in time, therefore it should be considered as a Transmission Function. In this paper, we continue with the work of analyzing the epidemic cycles, following the data treatment done in [1] and [2] and now applying the so-called Logistic Model to estimate three new parameters: the final count for the total infected, the daily infection rate and the lag time, which defines when the cycle really started. Additionally, we supply a novel model, based on the concept of inventory formation, in order to analyze and determine the actual incubation period for the virus.

## 2. Logistic Model

Considered by many authors as a good fit for modeling pandemic and epidemic episodes [3], the Logistic Model describes all three typical phases: the slow start, the steady growth and finally the asymptotically behavior at the end. There are several ways to describe this function and this work will use the so-called Richard Growth Model or Curve, to describe the accumulated number of infected cases in some countries. This formulation also allows for three parameters estimation: the final count for the total infected, the daily infection rate and the lag time. The generalized logistic function has the following form:

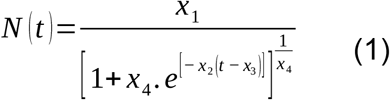

By selecting the highest r^2^ among several variations of (1), through curve-fitting, a particular form for (1) is found such as:

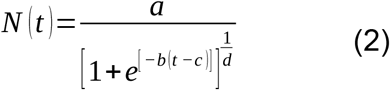

Where N(t) is the number of infected persons at a given period of time t, a is the final count for the total infected, b is the daily infection rate, c is the lag phase and d is a positive real number. It can be shown that:

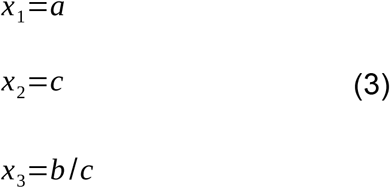

The constants a, b, c, and d will be used to estimate x_1_, the maximum number of infected people in a given place. X_2_ is the daily Infection Rate, or the average absolute daily increase in number of infected, which can be used to determine the Reproduction Number and to estimate the Incubation Period. Finally, x_3_ is used to estimate the Lag Phase, or the actual time when the first case happened.

## 3. Case Studies on the Number of Infected

In order to show numerical cases of the application of the proposed model, the data from three European countries with different closed cycles, in accordance to the criteria presented in [1], are analyzed: Germany, a country that was reported as exemplary in terms of application of Non-pharmaceutical Interventions (NPIs); Italy, which stayed at the center of the initial crisis; and Sweden, which didn’t apply strong NPIs in general. All data was collected from the John Hopkins University’s website (https://coronavirus.jhu.edu/map.html) on the COVID-19 on the declared dates. The determination of X_3_ allows to predict when the first case really occurred.

### 3.1 Germany

Data collected for Germany from February 15th to July 20th is plotted in Figure 1. The blue dots represent the daily registered infected cases submitted to MAMI, the red continuous line represents the Richard Growth Model curve, and drawn using parameters determined by the MAMI data.

**Figure 1–.**
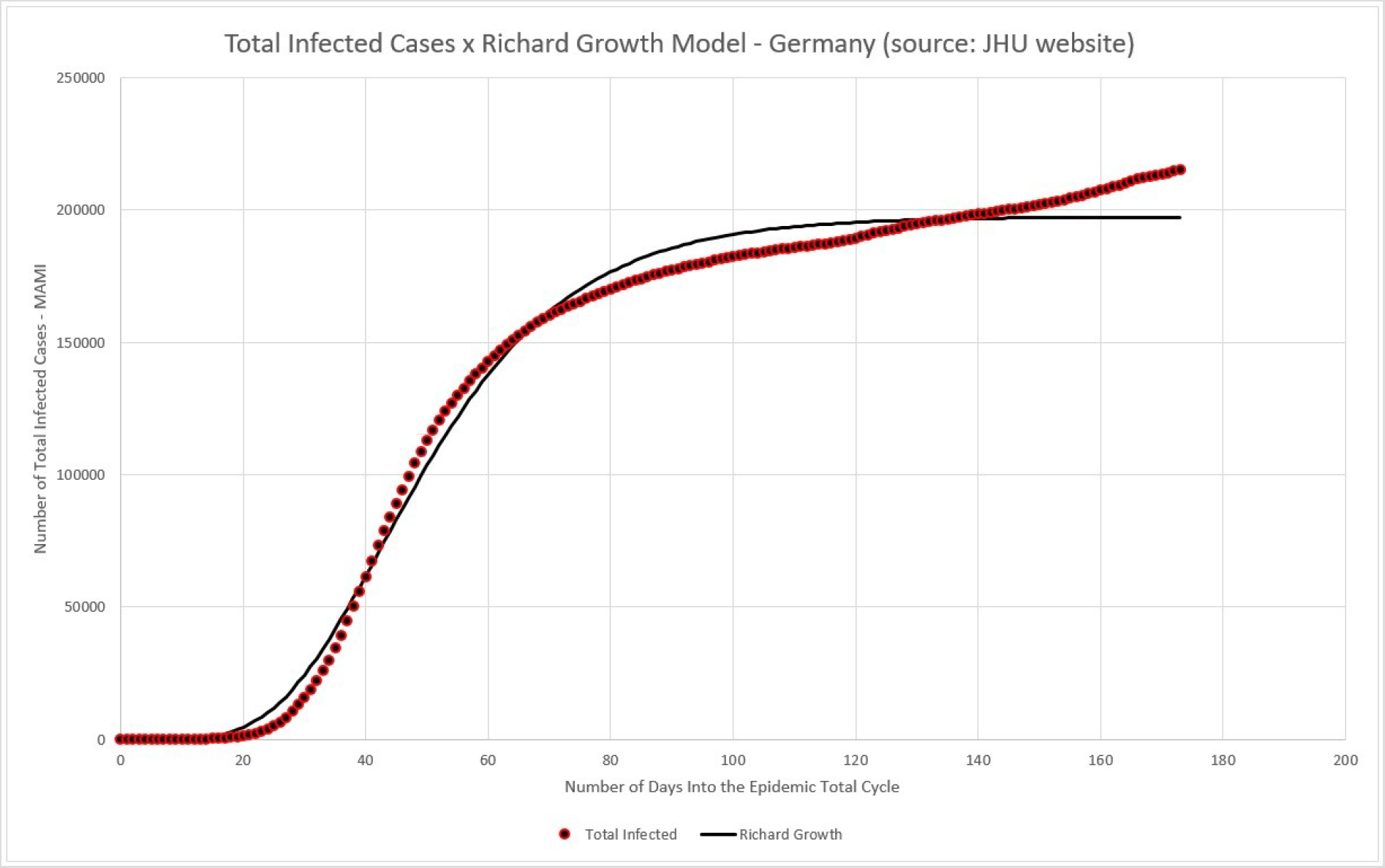
Germany – total number of infected (MAMI) compared to Richard Growth Model prediction.

As discussed in previous work [2], the German critical epidemic cycle started at March the 6^th^. According to Table 2, the first case must be recorded 89 days before that, with X_3_ indicating that the first case of the total epidemic cycle occurred around December, 8^st^ 2019.

**Table 1.** Curve-fitting data.

| Curve-Fitting |  |
| --- | --- |
| a | 197372.97 |
| b | -5.2260 |
| c | 0.0587 |
| d | $4.4208 \times 10^{-4}$ |

**Table 2.**
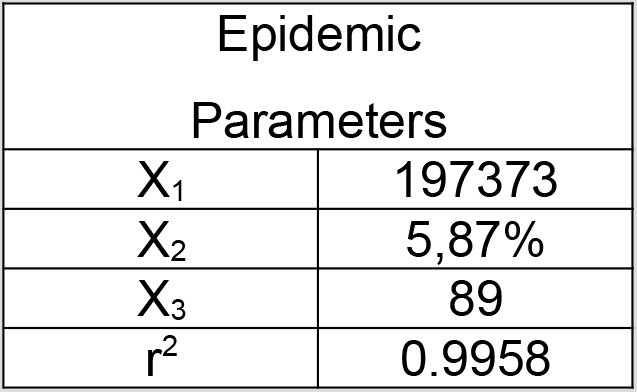
Epidemic parameter determined using curve-fitting data from Table 1.

| Epidemic Parameters |  |
| --- | --- |
| $X_1$ | 197373 |
| $X_2$ | 5,87% |
| $X_3$ | 89 |
| $r^2$ | 0.9958 |

### 3.2 Italy

Data collected for Italy from February 15th to July 20th is plotted in Figure 2. The blue dots represent the daily registered infected cases submitted to MAMI, the red continuous line represents the Richard Growth Model curve, and drawn using parameters determined by the MAMI data.

**Figure 2.**
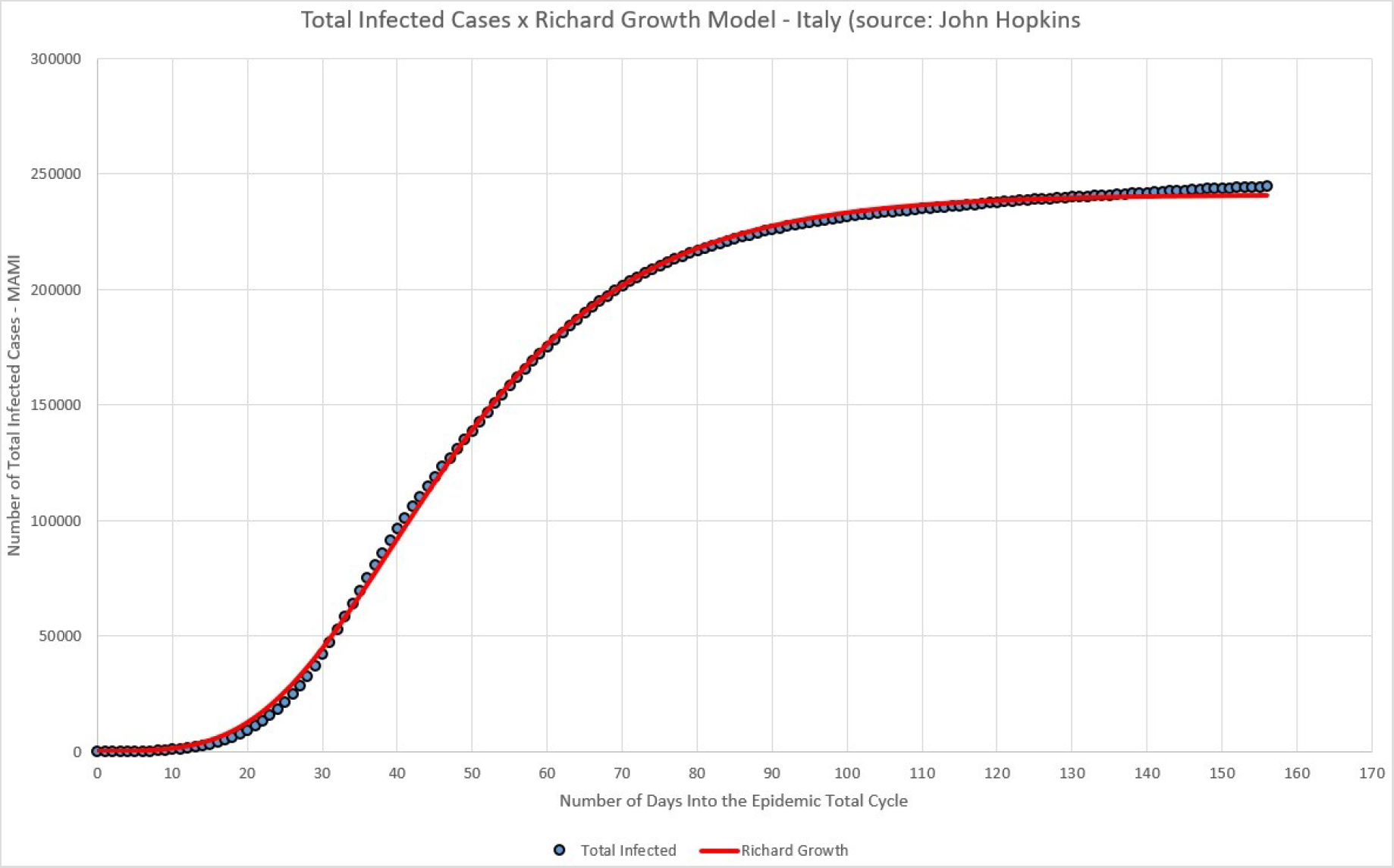
Italy – total number of infected (MAMI) compared to Richard Growth Model prediction.

According to [2], the Italian critical epidemic cycle started at February the 25^th^. According to Table 4, the first case must be recorded 86 days before that, with X_3_ indicating that the first case of the total epidemic cycle occurred around December, 1^st^ 2019.

**Table 3.**
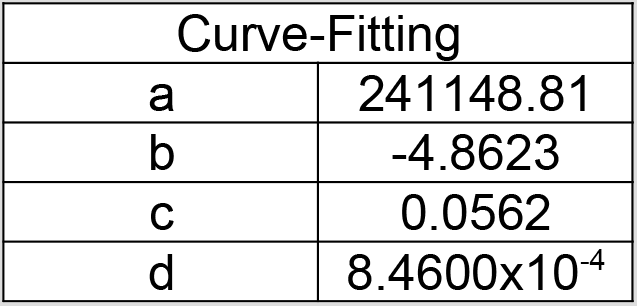
Curve-fitting data.

**Table 4.** Epidemic parameter determined using curve-fitting data from Table 3.

| Epidemic Parameters |  |
| --- | --- |
| $X_1$ | 241149 |
| $X_2$ | 5,62% |
| $X_3$ | 86 |
| $r^2$ | 0.9995 |

### 3.3 Sweden

Data collected for Sweden from February 15th to July 20th is plotted in Figure 3. The blue dots represent the daily registered infected cases submitted to MAMI, the red continuous line represents the Richard Growth Model curve, and drawn using parameters determined by the MAMI data.

**Figure 3.**
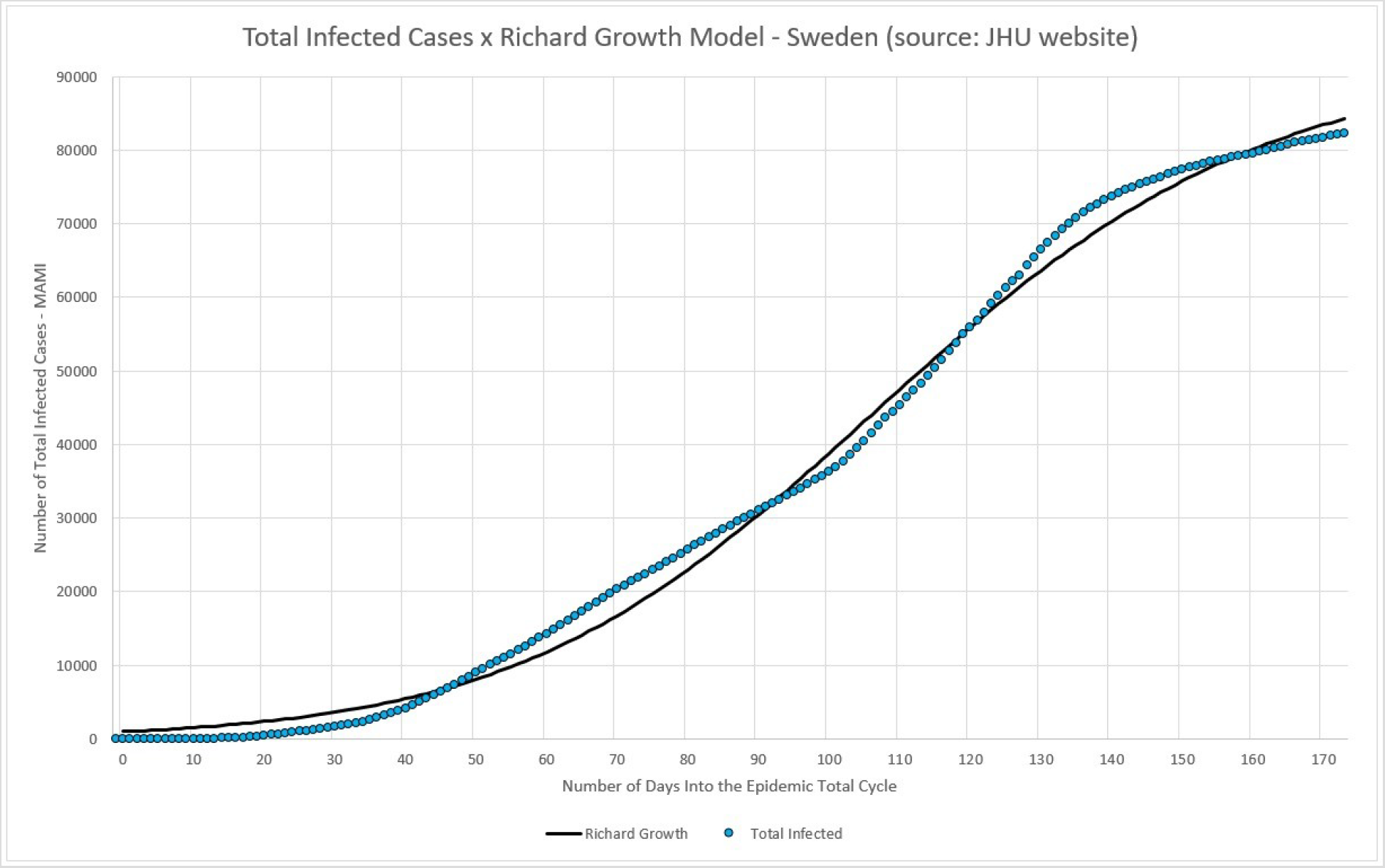
Sweden – total number of infected (MAMI) compared to Richard Growth Model prediction.

In [2] is shown that the Swedish critical epidemic cycle started at March the 4^th^. According to Table 6, the first case must be recorded 98 days before that, with X_3_ indicating that the first case of the total epidemic cycle occurred around November 27^th^ 2019.

**Table 5.** Curve-fitting data.

| Curve-Fitting |  |
| --- | --- |
| a | 92538,59 |
| b | 3.4050 |
| c | 0.0348 |
| d | $7.5514 \times 10^{-1}$ |

**Table 6.**
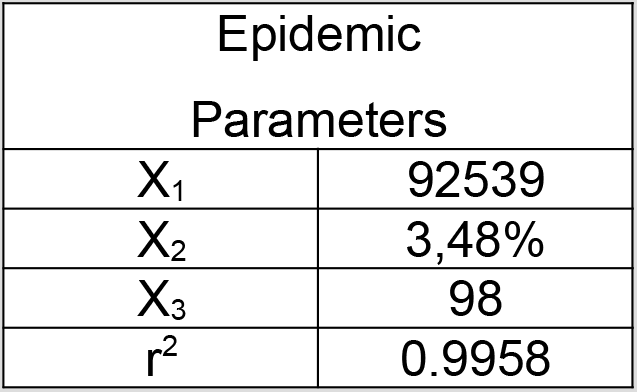
Epidemic parameter determined using curve-fitting data from Table 5.

## 4. Incubation Period Estimation

Although there are a series of studies on the incubation period for Sars-CoV-2, in order to maintain consistency with the rest of the modeling performed since [1] and [2], we seek to develop a model that could also estimate what would be the best incubation period estimation method to consider when modeling epidemic cycles. For that, we defined a model of stock of infected people similar to the productive systems, as shown in (4).

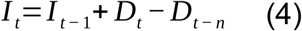

Where:

I_t_: Inventory of people infected in day t, or the total of infected in day t;

I_t-1_: Inventory of people infected in the previous day;

D_t_: Number of people detected with the disease in day t;

D_t-n_: Number of people detected with the disease n days before t;

The equation described in (4) should be interpreted as follows: the number of people that The number of people who are infectious on a given day is equal to the number of people who were infectious the day before, plus the number of infectives detected on the same day, minus the number of people who have left the N-day incubation period. This reasoning therefore assumes that as soon as a person finds out he/she is infected, that is, when this person leaves the incubation period, assumes perfect isolation and stops infecting. Although this assumption is not perfectly realistic, since it depends not only on individual responsibility, but also on the implementation of isolation measures, at the same time it must also be considered that not every infected person effectively infects others, since, knowing that the isolation is not the only way to avoid contamination by viruses, there are cases, for example, you may have people in your social life immunized in some way. Thus, we consider this assumption to be reasonable to be applied statistically.

Other basic assumptions is that of all people able to be contaminated (not vaccinated, exposed enough to the pathogen, etc…) not all of them will develop the disease in a form severe enough to be noticed. So, the recorded number of daily cases reflect not the total contaminated, but those who seek medical attention and therefore were diagnosed as contaminated.

Therefore, It is the number of infectives in a given day, or the “inventory” of people that can infect other people in a given day. With the formulation defined in (4) and the assumptions described previously, we followed with the analysis and simulations for the three countries.

### 4.1 Case Studies of the Number of Infectives

In this topic we approach the model of infectious inventory for the three countries considered. Simulations are made for incubation cycles of 3, 5, 7, 9 and 11 days. Inventories are calculated according to the formula presented in (4) and plotted together with the MAMI of detected cases.

One cannot take the assumptions that come together with formula (4) as deterministic, nor consider that formula (4) describes a perfect “production” system. There is no biological system that behaves in such perfect and deterministic way. Therefore, the data shown in Figures 4, 5, and 6 are not conclusive by themselves, because the imperfections of the contamination paths, or the “production system”, should be considered.

**Figure 4.**
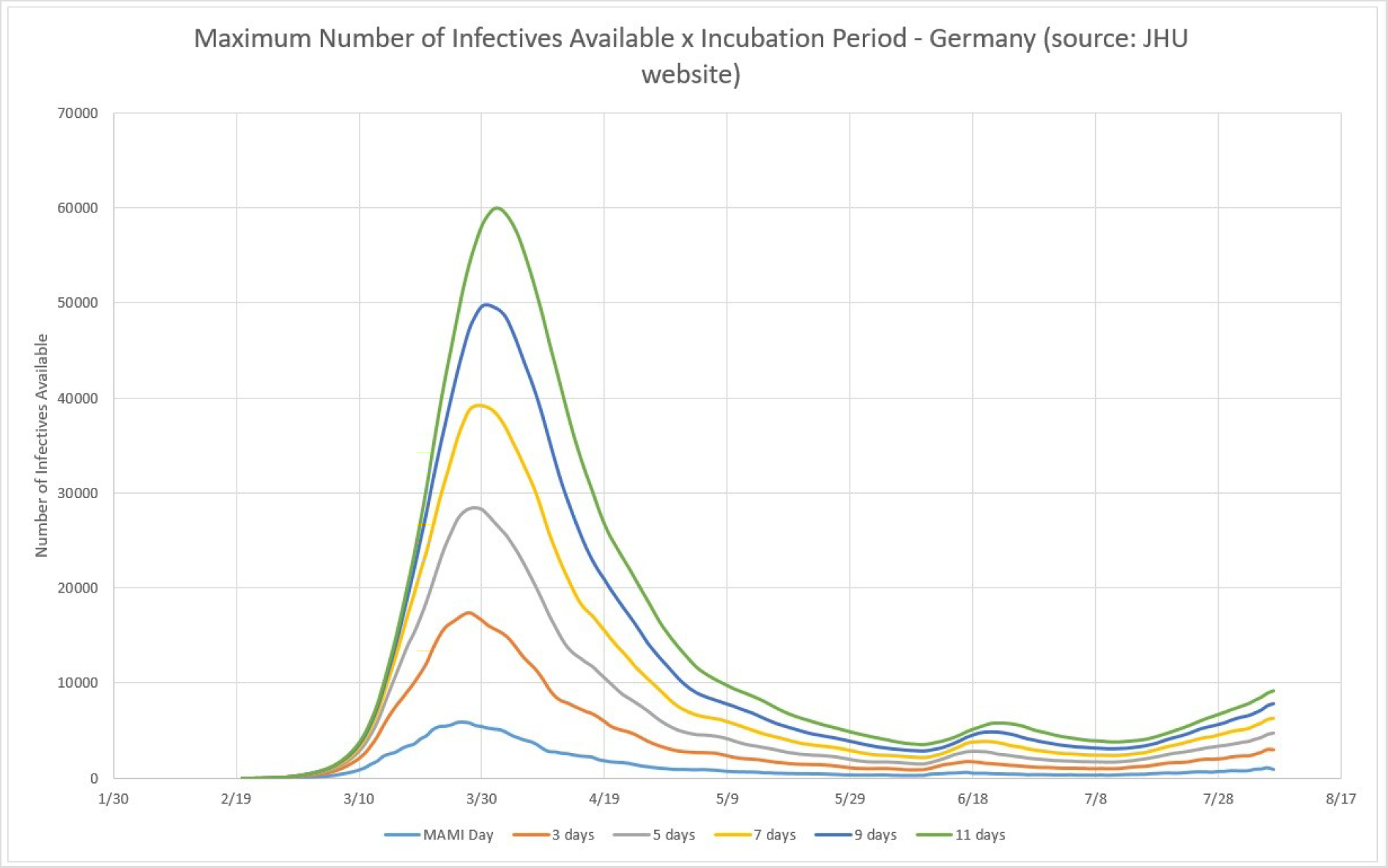
Germany: Infectives Inventories for 3, 5, 7, 9, and 11 days of incubation, compared to MAMI.

**Figure 5.**
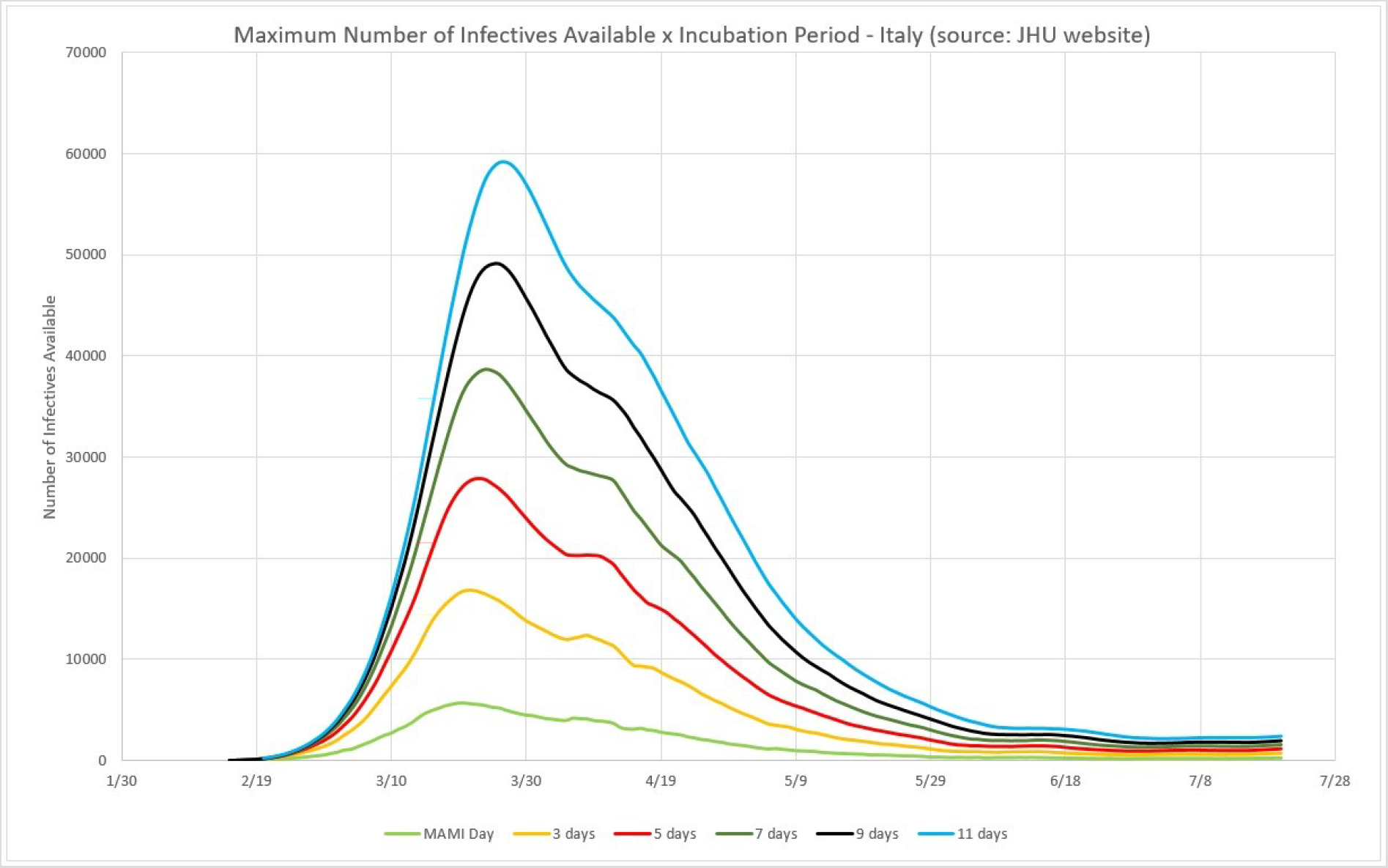
Italy: Infectives Inventories for 3, 5, 7, 9, and 11 days of incubation, compared to MAMI.

**Figure 6.**
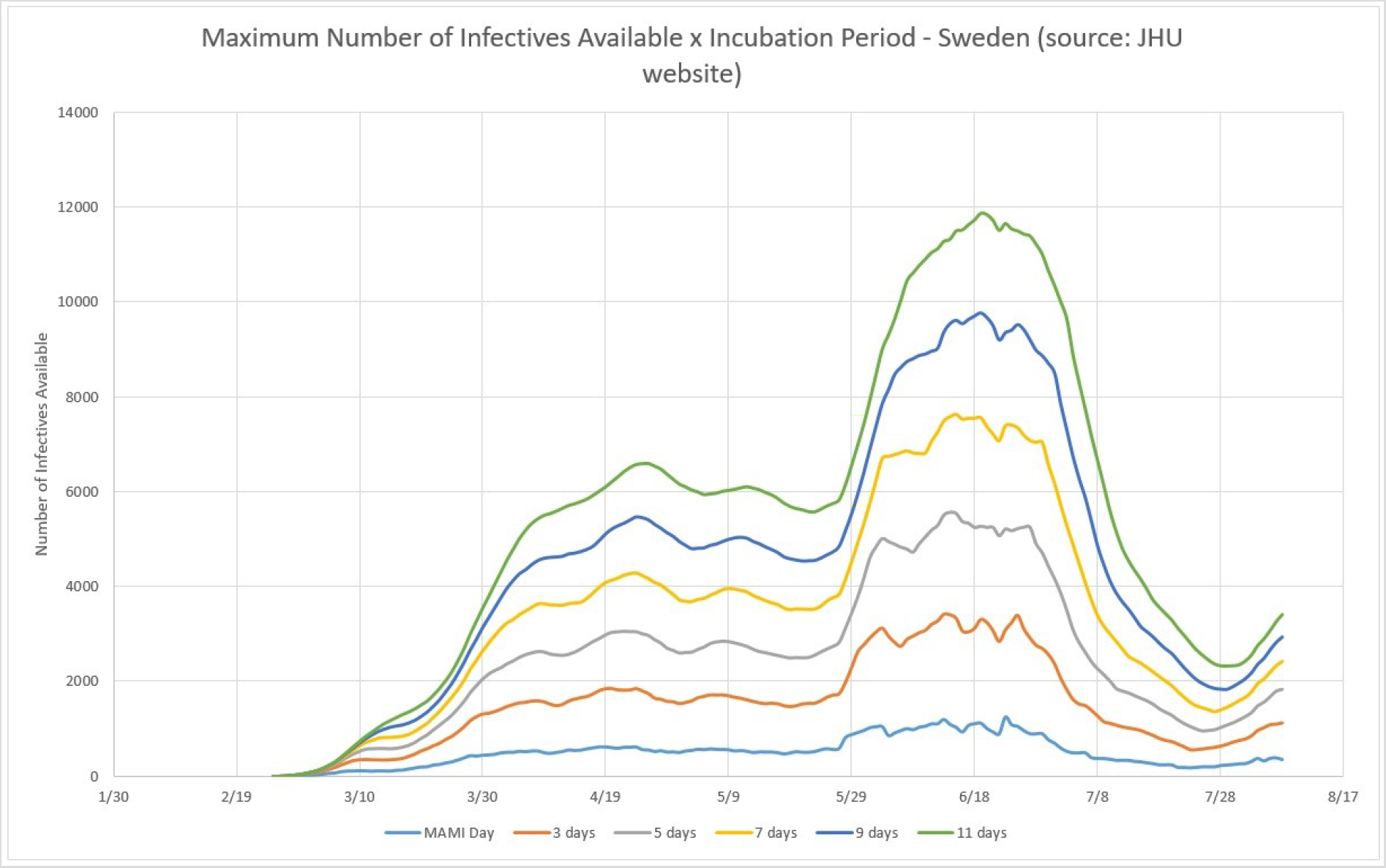
Sweden: Infectives Inventories for 3, 5, 7, 9, and 11 days of incubation, compared to MAMI.

### 4.3 Efficiency of the Infection System

According to the experimental data obtained, the efficiency of the biological system here described – the COVID-19 pandemic – has a Power function form, as shown in Figure 7. Although the three countries here analyzed have very different epidemic cycles, the percentage of people infected compared to the incubation period varies very little. This reflects, probably, the fact that the incubation period is in fact a constant value. Figure 7 shows that, for example, for a 5-day incubation period, the percentage of people who were exposed to the virus and displayed symptoms severe enough to prompt them to look for medical care, was around 20%. In other extreme, if the virus had a 11-day incubation period, the numbers of actual cases registered would have indicated a 10% rate of infection in the general population.

This curve, although restricted to only the three countries, covers nations with quite different NPI policies, population sizes, and land masses. It shows that, according to registered cases, Sars-CoV-2 affects a small segment of these populations and at the same proportions. The sub-notification effect does not interfere with this curve behavior, once unregistered cases only happen into the complementary percentage, which goes without symptoms, which confirms the findings in [2].

**Figure 7.**
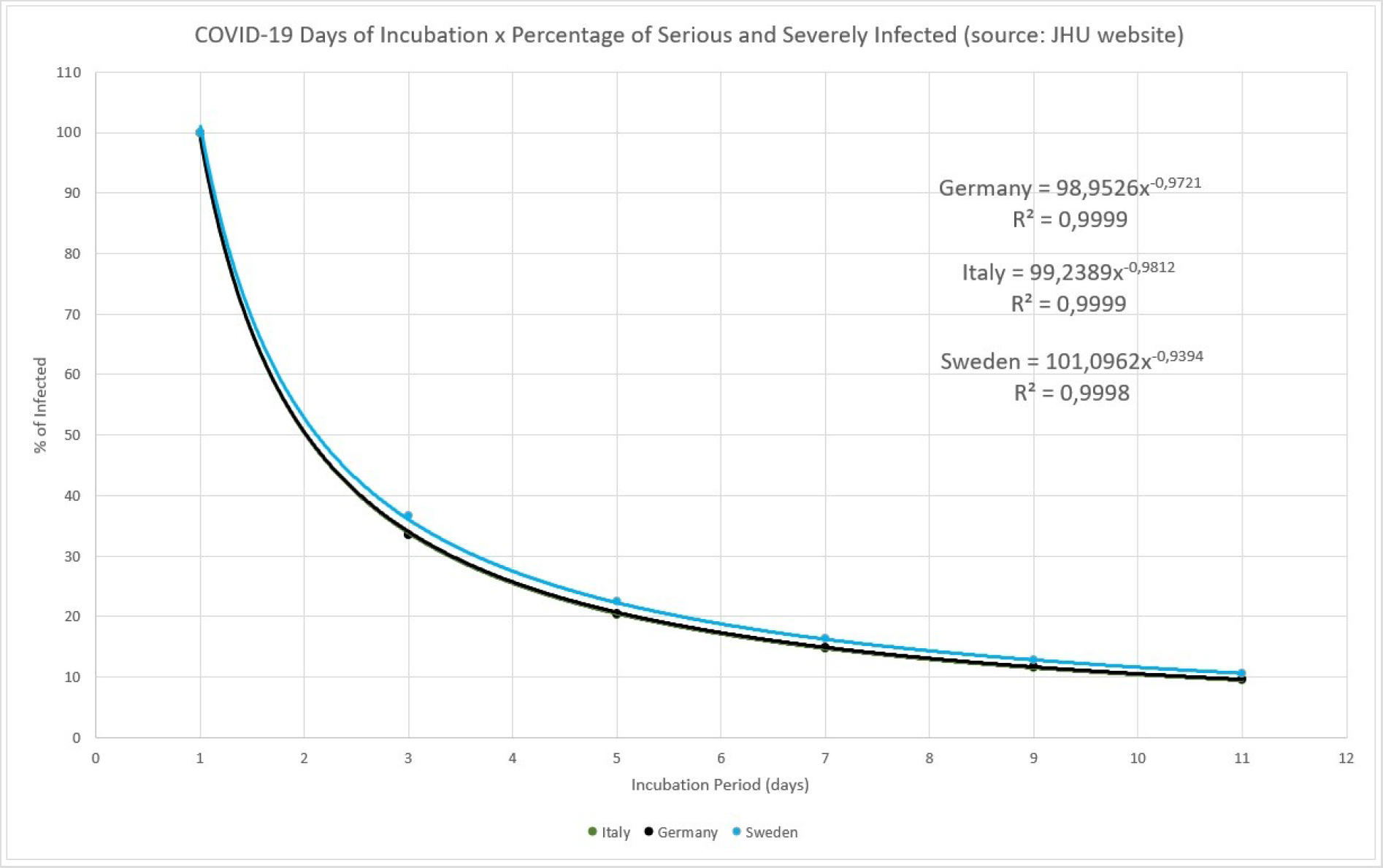
COVID-19 Number of days of incubation versus percentage of serious and severe infected.

The conclusion, putting together formula (4) with the efficiency measurement in Figure 7, is that the reported rate of 80% of sub-notification [4], or 20% of people with more serious symptoms, represents 1/5 of the Inventory of the Infective persons. In other words, there is five times more persons in infective state than the detected and reported by the MAMI numbers, leading to a 5-day of incubation period.

### 4.4. Sub-Notification Estimation

Here it is understood that sub-notification is referred to the amount of people exposed and contaminated by the Sars-CoV-2 and that never had displayed any symptoms or they were so mild that the bearer chose to ignore, therefore never being part of the actual registered daily cases. It is the complement to percentage discussed in the previous item, divided by the actual registered number. The estimation is quite simple: given the Incubation Period, how many times the registered amount should be multiplied to correctly express the estimated sub-notification? For example, for a 5-day incubation period in Germany, it is expected a sub-notification around 4 times the registered number of cases in any given day. If 100 were registered, 400 were not.

**Figure 8.**
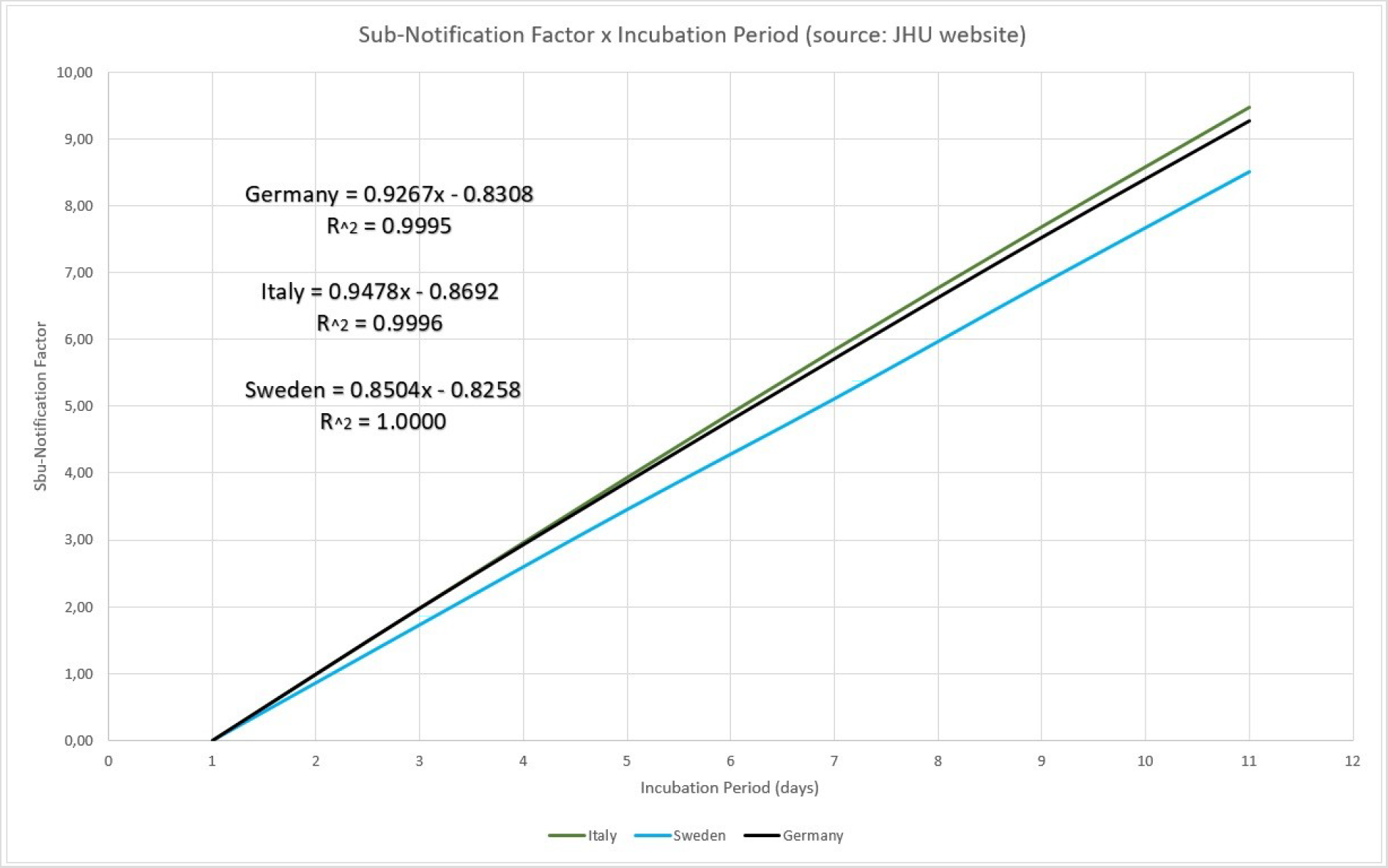
Sub-notification factor.

## Conclusions

There is the awareness that local epidemic COVID-19 cycles may have started earlier, without proper identification. Starting dates influence all the parameters that govern the statistical models used for controlling the infection, therefore, this work proposed two models based on experimental data. The Logistic Model was used to obtain three parameters of the epidemic cycle of COVID-19, the number of total infected, the daily infection rate and the lag phase, which determined the actual probable onset of the epidemic for the studied countries – solving the problem of “contamination” of the other parameters by wrongly determined onset dates.

Complimentary, a novel model based on the concept of “inventory of infective persons” is used to calculate the number of infective persons, as well as to determine numerically the incubation period, proving that it is most probable that is 5-day long.

## Data Availability

All data was collected from the John Hopkins University website (https://coronavirus.jhu.edu/map.html) on the COVID-19 on the declared dates.

## References

1. De Carvalho EA, De Carvalho RA. Identification of Patterns in Epidemic Cycles and Methods for Estimating Their Duration: COVID-19 Case Study. MedRxiv (2020). Published 2020 Jul 15. doi: https://doi.org/10.1101/2020.07.13.20153080

2. De Carvalho E. A., De Carvalho R. A. COVID-19: Time-Dependent Effective Reproduction Number and Sub-notification Effect Estimation Modeling. MedRxiv (2020). Published 2020 Jul 28. doi: https://doi.org/10.1101/2020.07.28.20164087

3. Hilbe J. M., Logistic Regression Models. Chapman & Hall/CRC Texts in Statistical Science, 1st Edition, 2017.

4. World Health Organization (WHO). Similarities and Differences Between COVID-19 and Influenza. https://www.who.int/emergencies/diseases/novel-coronavirus-2019/question-and-answers-hub/q-a-detail/q-a-similarities-and-differences-covid-19-and-influenza. Accessed on 23th of July, 2020.

